# Most clinical trials involving American children that violated FDAAA legal reporting requirements had not published outcomes in the scientific literature

**DOI:** 10.1101/2023.01.17.23284683

**Authors:** Till Bruckner, Samruddhi Yerunkar, Okan Basegmez, Ronak Borana, Belén Chavarría, Mayra Velarde, Nicholas J. DeVito

## Abstract

Non-publication, incomplete publication and excessively slow publication of clinical trial outcomes contribute to research waste and can harm patients. In this cohort study, we used public ClinicalTrials.gov registry records to identify 81 paediatric clinical trials in the United States that appear to have violated the FDA Amendments Act 2007 (FDAAA) reporting requirements. We then searched the literature for the outcomes of these 81 trials and contacted trial sponsors about the status of results.

We found that only 22/81 trials (27.2%) had made their results public in a full-length peer-reviewed publication, and that only 8/81 (9.9%) trials had done so within one year of their primary completion date. Our findings highlight the need for US Food and Drug Administration and the National Institutes of Health to systematically monitor FDAAA compliance and enforce reporting requirements.

## Background

Non-publication of clinical trial results is unethical^1^ as it leaves gaps in the medical evidence base that can harm patients and contributes to research waste.^2,3^ Delays in making clinical trial results public can hamper the development and adoption of new or improved medical interventions. Trial registries offer a way to audit widespread shortcomings in the reporting of trial outcomes in the academic literature.^2,4,5 6,7^

The Food and Drug Administration Amendments Act of 2007 (FDAAA) set out to curb reporting biases and accelerate results sharing in the public domain. Entities sponsoring certain types of clinical trials are legally obliged to make the tabular summary results of those trials available on the public ClinicalTrials.gov trial registry within one year of a trial’s primary completion date.^8^

Enforcement of the law’s provisions by the U.S. Food and Drug Administration (FDA) has so far been lacklustre acting only on a tiny subset of all delinquent trial results since the passage of FDAAA.^9^ Despite a 2016 promise by U.S. president Joe Biden to cut off public funding to institutions that violate the law,^10^ the National Institutes of Health have so far not done so, and have been reprimanded by the Office of the Inspector General for failing to ensure funded research has been reported in a timely manner.^11^

Previous research has shown that while sponsor compliance with FDAAA has improved in recent years, not all sponsors consistently meet their legal obligations, and thousands of results for such trials remain missing from the registry.^12^ A recent study found that nearly half (49.8%) of completed paediatric trials registered on ClinicalTrials.gov between 2007 and 2020 were unreported in either the literature or on ClinicalTrials.gov.^13^ Earlier, a study had found significant publication bias in This is consistent with prior work suggesting publication bias in smaller samples of paediatric trials.^14–21^ However, there has been no prior research comprehensively assessing the nature and reporting behaviour of paediatric trials in apparent violation of the FDAAA.

We therefore set out to assess the publication status and publication speed of paediatric trials that appear, based on registry data, to be subject to FDAAA reporting requirements, and as of September 2022 had failed to make their results public on ClinicalTrials.gov as required by law.

## Methods

This study was preregistered on OSF (https://osf.io/me9q8). A UK Health Research Authority NHS REC ethics waiver was secured on 20 October 2022. The study protocol, dataset, correspondence with FDA and sponsors, literature search guide and ethics waiver are publicly available on GitHub (https://github.com/TillBruckner/FDAAApaediatric). The outcomes of this study are reported in line with the STROBE guideline for cohort studies. This study did not receive external funding.

### Data Acquisition & Cohort Generation

A researcher (NJD) downloaded a dataset of all clinical trials registered on ClinicalTrials.gov as of 08 September 2022 and flagged 34,649 trials that appear subject to FDAAA based on registered data elements using a previously published methodology.^12^ The population of FDAAA trials was then narrowed down, prior to study commencement, by another researcher (TB). Trials with a maximum age of 18 or above or stated maximum age, trials that were not yet due to report results (i.e., a primary completion date <1 year ago), trials that had results available on the registry, trials with results pending, trials with a delay certificate, and trials whose location data excluded the United States were removed to form the cohort. This left 97 trials, of which 16 stated no location data. Out of those 16 trials, 13 were excluded because their sponsor was located outside the United States. This process yielded a cohort of 84 clinical trials that exclusively involved children, included study sites in the United States, and were in apparent violation of FDAAA reporting requirements.

### Search Strategy

A different researcher (SY) performed a systematic registry and literature search for the results of these 84 trials in late October 2022. The search involved: 1) checking ClinicalTrials.gov records for native tabular results (submitted after 8 Sept 2022) and any linked journal articles either uploaded by the sponsor or automatically linked to the registry; 2) searching Google Scholar for the ClinicalTrials.gov NCT ID; and 3) searching Google Scholar using the trial title and listed primary investigators. Another team member (TB) then reviewed the data and followed up on selected trials, including all that had been marked as difficult to classify. One trial (NCT01072370) was eliminated from the cohort as the sponsor had changed its status after data extraction from “terminated” to “withdrawn”, indicating the trial never actually took place, leaving 83 eligible trials.

### Stakeholder Outreach

We emailed the FDA press office a list of all 83 trials requesting information on related FDA enforcement actions in November 2022, followed by a reminder email; FDA never responded. In parallel, two researchers (SY,TB) in November contacted the sponsors of all trials for which we had been unable to locate outcomes on the registry or in full-length peer-reviewed publicationsby email to check whether relevant publications had been overlooked. We used sponsors’ press offices as point of contact whenever possible. In the case of 4 investigator-initiated trials (IITs); we contacted the institution associated with the primary investigator. In the case of companies that had merged or been acquired, we extracted contact details for the respective successor companies, and listed them as the responsible sponsors within our spreadsheet. We were unable to identify contact emails for five commercial sponsors responsible for sox trials. Three sponsor companies had ceased to exist and no successor entity could be identified: HeadSense Medical (1 trial), Lachlan Pharma Holdings (1 trial), Micelle BioPharma Inc (2 trials). Two sponsors had no contact information on their websites: Pharmaceutical Project Solutions (1 trial) and Molex Ventures (1 trial) and could not be reached via social media. In December, we sentreminder emails to contactable sponsors who had not responded to our initial outreach.

In total, 13 of 48 (27.1%) unique sponsors we contacted responded. Two sponsors informed us that their trials were still ongoing and that the primary completion dates (PCD) stated in the registry were thus incorrect (Gillette Children’s Specialty Healthcare, NCT03107546 (PCD now updated); Carilion Clinic, NCT03364218 (as of writing, still displays incorrect PCD)). We removed those 2 trials from the cohort, leaving 81 trials. One sponsor flagged a publication that our initial literature search had missed; this was included in our dataset. The other sponsors did not flag salient published results.

### Secondary Searches

In December 2022, four team members (RB, BC, MV, OB) conducted a second systematic literature search using PubMed, covering all trials for which a full journal publication had still not been located. They located publications containing the results of 2 additional trials (NCT03937869 and NCT04560478). On 24 December 2022, one team member (TB) performed a final search of ClinicalTrials.gov for tabular summary results for all 81 trials as a final follow-up on the results availability of the cohort on ClinicalTrials.gov.

### Outcomes

As per study protocol, we segmented trials into those with tabular summary results available on ClinicalTrials.gov, and those still missing results in apparent violation of FDAAA. For the latter cohort, we documented whether and when they had made their results public in journals. For all trials, we tabulated the number of enrolled children using ClinicalTrials.gov data as of 08 September 2022.

## Results

Our final cohort contained 81 clinical trials that exclusively involved children, and that included children located in the United States, and that appeared to be in violation of the legal requirement to make their results public within 12 months of their primary completion date under the FDAAA 2007.

### Results Availability

Only 22/81 trials (27.2%) had made their results public in a full-length peer-reviewed publication at that point in time. A further 5/81 trials (6.2%) had made their results public only partially or in a non-journal format; these are classified as not reported below. The remaining 54/81 trials (66.7%) remained completely unpublished.

Publication timelines for 14/22 journal articles (63.6%) exceeded one year post primary completion date.

**Table 1:** Publication speed and research waste in paediatric trials missing CT-gov results, # of trials

|  | Trials total | Timely publication | Delayed publication |  |  | Research waste |
| --- | --- | --- | --- | --- | --- | --- |
|  |  | Within 1 year* | Within 1-2 years | Within 2-3 years | After 3 years | No results |
| **All trials** | 81 | 8 | 11 | 2 | 1 | 59 |

\* Including NCT02010034 & NCT02102698

Overall, less than 10% of paediatric trials that were in violation of FDAAA reporting requirements had made their results public in journals within a year of their primary completion dates.

**Table 2:** Publication speed and research waste in paediatric trials missing CT-gov results, % of trials

|  | Timely publication | Delayed publication |  |  | Research waste* | Publication speed (days, average)** |
| --- | --- | --- | --- | --- | --- | --- |
|  | Within 1 year | Within 1-2 years | Within 2-3 years | After 3 years | No results |  |
| **All trials** | 9.9% | 13.6% | 2.5% | 1.2% | 72.8% | 501 |

\* Including partial and non-journal publications
\*\* Excluding NCT02010034 & NCT02102698

In total, 5,803 children participated in the 81 clinical trials that were in violation of FDAAA reporting requirements per their ClinicalTrials.gov registry data. As the six trials in our cohort with international study arms only enrolled a total of 72 participants, virtually all children were recruited at U.S. study sites. The outcomes for 4,408 paediatric trial participants had not been made public in peer-reviewed journals. Results for only 410 participants (7.1%) had been made public in the peer-reviewed literature within a year of trial completion.

**Table 3:**
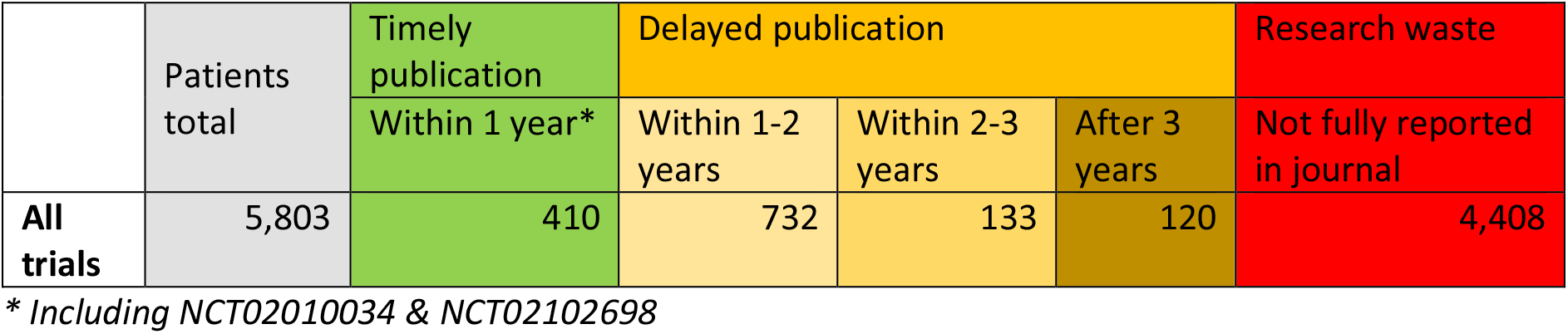
Publication speed and research waste in paediatric trials missing CT-gov results, # of patients

By 24 December 2022, tabular summary results had been uploaded for 16/81 trials onto ClinicalTrials.gov, 14 of which had been sponsored by entities contacted by the project team. Of note, the sponsor Benuvia Therapeutics did not respond to outreach but had uploaded the results of 4 trials.

However, as of the final follow-up in December, a total of 43 clinical trials involving 3,627 children remained completely unreported both in the academic literature and on ClinicalTrials.gov.

## Discussion

### Summary of Findings

We identified 81 clinical trials involving 5,803 children that were in violation of FDAAA reporting requirements as they had failed to upload results to ClinicalTrials.gov within a year of primary completion. For the majority of these trials, reporting to ClinicalTrials.gov would have been the primary source of results as just 22 (27.2%) trials had results that could be identified in the literature. The majority of trials with results (63.6%) had results appear in the literature in excess of the one year reporting deadline required in the FDAAA. While reporting to ClinicalTrials.gov improved upon followup, at the completion of study follow-up in December 2022, 43 paediatric trials, with a planned or actual enrolment of 3,627 children had not made results public on ClinicalTrials.gov or in the literature.

### Strengths and limitations

This study provides a systematic overview of the results status of paediatric trials apparently in violation of FDAAA Section 801 reporting requirements. The study was pre-registered and the underlying data made public. Data quality was safeguarded through two independent rounds of structured literature searches combined with direct outreach to sponsors.

By design, our cohort was limited to clinical trials exclusively involving U.S. children that were identifiable as such from registry data. This excluded trials that set out to recruit both adults and children, and trials for which registry data did not state maximum participant ages. Importantly, our cohort was further limited to trials completed since 2017 that fall under the purview of the FDAAA Final Rule, a regulatory document that clarified various aspects of FDAAA requirements and allowed for improved independent identification and audit of the law’s requirements. As such, we did not capture older trials dating back to the law’s passage in 2007 which per a 2020 court ruling are also subject to FDAAA reporting requirements when testing an FDA-approved intervention.^22^ Therefore, there are almost certainly numerous additional trials involving U.S. paediatric participants that remain unreported in violation of FDAAA as well as unavailable in the literature. We also excluded paediatric trials that, while subject to FDAAA, did not involve U.S. study sites even though some non-US trials are covered under the FDAAA in certain circumstances. Registry data for some trials stated enrolment targets rather than actual enrolment. Although we aimed to validate our sample with sponsors, we received confirmation on the reporting status for only 15/52 trials (28.8%) that we had flagged with sponsors.

Our outreach appears to have motivated multiple sponsors to rapidly upload overdue results onto ClinicalTrials.gov. However, this is an explorative finding as this outreach was done to validate literature search results; this study was not designed to test the impact of outreach on reporting behaviour.

## Conclusion

In total, 43 trials involving 3,627 children identified in this study remained completely unreported. Trial participants included children suffering from life-threatening conditions, including congenital heart disease (NCT04040452) and Duchenne Muscular Dystrophy (NCT03917719). One trial involved prematurely born babies with a birth weight of less than 1,000 grammes (NCT02524249).

Our findings highlight the urgent need for FDA and the National Institutes of Health to systematically enforce FDAAA reporting requirements in order to curb research waste, reduce publication bias, and accelerate medical progress.

## Supporting information

Dataset of 81 FDAAA paediatric trials

## Data Availability

All data produced are available online at: https://github.com/TillBruckner/FDAAApaediatric

